# Prevalence and consequences of *APC* mosaicism in patients with colorectal adenomas

**DOI:** 10.1101/2025.02.18.25322465

**Authors:** Diantha Terlouw, Manon Suerink, Yentl Buitelaar, Monique E van Leerdam, Frederik J Hes, Demi van Egmond, Dina Ruano, Anja Wagner, Floris H Groenendijk, Isabelle C Meijssen, Eline Overwater, Sanne W Bajwa-ten Broeke, Arjen R Mensenkamp, Iris D Nagtegaal, Carli M Tops, Alexandra M J Langers, Tom van Wezel, Hans Morreau, Maartje Nielsen

**Affiliations:** Department of Pathology, Leiden University Medical Center, Leiden, the Netherlands; Department of Clinical Genetics, Leiden University Medical Center, Leiden, the Netherlands; Department of Gastroenterology and Hepatology, Leiden University Medical Center, Leiden, the Netherlands; Vrije Universiteit Brussel (VUB), Universitair Ziekenhuis Brussel (UZ Brussel), Clinical Sciences, research group Reproduction and Genetics, Centre for Medical Genetics, Universitair Ziekenhuis Brussel (UZ Brussel), Brussels, Belgium; Department of Clinical Genetics, Erasmus University Medical Center, Rotterdam, the Netherlands; Department of Pathology, Erasmus MC Cancer Institute, Rotterdam, the Netherlands; Department of Genetics, University Medical Center Groningen, Groningen, the Netherlands; Department of Human Genetics, Radboud University Medical Centre, Nijmegen, the Netherlands; Department of Pathology, Radboud University Medical Centre, Nijmegen, the Netherlands

**Author notes:** **Corresponding author** Dr. M. Nielsen, Department of Clinical Genetics, Leiden University Medical Center, Albinusdreef 2, 2333 ZA Leiden, The Netherlands. shared last author (Tom van Wezel, Hans Morreau, Maartje Nielsen). **Author contributions** DT: acquisition of data, analysis, and interpretation of data, drafting the manuscript. MS: acquisition of patients, interpretation of data, critical revision of manuscript. YB: acquisition of data, critical revision of manuscript. MEvL: acquisition of patients, critical revision of manuscript. FJH: acquisition of patients, critical revision of manuscript. DvE: lab support, critical revision of manuscript. DR: bioinformatical support, critical revision of manuscript. AW: acquisition of patients, critical revision of manuscript. FHG: acquisition of patients, critical revision of manuscript. ICM: acquisition of patients, critical revision of manuscript. EO: acquisition of patients, critical revision of manuscript. SWBtB: acquisition of patients, critical revision of manuscript. ARM: acquisition of patients, critical revision of manuscript. IDN: acquisition of patients, critical revision of manuscript. CMT: acquisition of data, critical revision of manuscript. AMJL: acquisition of patients, interpretation of data, critical revision of manuscript. TvW: study concept and design, interpretation of data, critical revision of manuscript. HM: study concept and design, acquisition of patients, interpretation of data, critical revision of manuscript. MN: study concept and design, acquisition of patients, interpretation of data, critical revision of manuscript.

**Keywords:** Colorectal adenomas, *APC*, mosaicism, Familial Adenomatous Polyposis

## Abstract

**Background and aims:** A substantial proportion of patients with adenomatous polyposis have no germline pathogenic variant in *APC*. The aim of this study was to determine the prevalence of *APC* mosaicism in these patients with unexplained polyposis and to draft guidelines for *APC* mosaicism testing and surveillance.

**Methods:** *APC* mosaicism was analyzed by targeted Next-Generation sequencing in 542 patients with a broad spectrum of polyposis phenotypes.

**Results:** The rate of *APC* mosaicism was 9.4%. This rate was 14.3% (46/322) in patients who meet the scope of national hereditary polyposis testing guidelines (≥10 adenomas before the age of 60 or with ≥20 adenomas before the age of 70). In patients who do not meet the scope of national guidelines, the detection rate was 2.3% (5/219). In patients with ≥20 adenomas before the age of 60, or ≥30 adenomas before the age of 70 the detection rate was ≥10%. Of 34 mosaic patients who underwent an esophagogastroduodenoscopy, 26% were diagnosed with gastroduodenal polyps. In one patient, the mosaic variant was detected in semen, but none of the children tested in this cohort inherited the mosaic variant.

**Conclusion:** We recommend *APC* mosaicism testing at least in patients negative for germline pathogenic variants with (1) ≥20 adenomas before the age of 60 or (2) ≥30 adenomas before the age of 70. Regular colonoscopy and at least one gastroduodenoscopy should be offered to *APC* mosaics, with frequency of follow-up based on findings. Offering germline testing for offspring should be considered.

## Introduction

Familial Adenomatous Polyposis (FAP), the most common polyposis syndrome, is caused by a pathogenic germline variant in the *APC* gene.^1^ FAP patients classically develop hundreds to thousands of colorectal adenomas. The severity depends on the genotype, with variants in the 5’ or 3’ end causing an attenuated phenotype with less than 100 colorectal adenomas (AFAP).^2^ In addition to colorectal adenomas, (A)FAP is associated with extracolonic manifestations such as duodenal and gastric neoplasms, osteomas, and desmoid tumors.^3, 4^ The detection rate of germline *APC* pathogenic variants is highly dependent on the number of adenomas, ranging from 1% in patients with 10-20 adenomas to 70% in patients with more than 100 adenomas in a large Dutch cohort.^5^ The detection rate of the second most common form of polyposis, *MUTYH*-associated polyposis (MAP), which has a recessive inheritance, ranges from 1% to 18% in the same cohort. However, a substantial proportion of adenomatous polyposis patients remain unexplained after routine diagnostic germline testing for *APC, MUTYH* and other polyposis associated genes like *NTHL1, POLD1* and *POLE*.

In 4% to 11% of unexplained polyposis patients, a so-called *APC* mosaicism can be identified analyzing leukocyte DNA.^6-8^ *APC* mosaics have a pathogenic *APC* variant in only a subset of body cells due to a *de novo* variant arising during embryogenesis.^9^ Depending on the timing of this occurrence, multiple or a single tissues are affected. Of note, studies that performed sequencing analysis on DNA isolated from colorectal lesions identified *APC* mosaicism in a much higher proportion, namely 25- 50%.^10-12^ These findings suggest a high prevalence of *APC* mosaicism in patients with unexplained polyposis that is not detectable in leukocyte DNA using standard diagnostic testing.

In general, the phenotypic characteristics of mosaics are milder than those observed in germline pathogenic variant carriers. This assumption suggests that *APC* mosaicism may also be present in less severe phenotypes such as 10-20 colorectal adenomas at the age of 60-70 years or with more than 20 adenomas at the age of over 70 years. These patients do not fall within the scope of the national hereditary polyposis testing guidelines and are, therefore, usually not tested.^13-15^

In this study, we performed targeted Next-Generation Sequencing (NGS) on DNA isolated from colorectal adenomas and/or carcinomas from patients with a broad spectrum of colorectal adenoma phenotypes. Based on the results, we aimed to draft guidelines for *APC* mosaicism testing and surveillance.

## Materials and methods

### Cohort description

*APC* mosaicism testing was performed on 541 patients from three medical centers in the Netherlands. The majority, 462 patients, were tested at the Leiden University Medical Center (LUMC), 44 at the Erasmus Medical Center (EMC) in Rotterdam and 35 patients from the University Medical Center Groningen (UMCG) were tested at the Radboud university medical center (Radboudumc).

#### Leiden (LUMC)

All patients who met the testing guidelines (either ≥10 cumulative adenomas before the age of 60 years or ≥20 adenomas between the ages of 60 and 70 years) were tested for germline variants at the Department of Clinical Genetics. Patients who did not meet the guidelines were included if they fell into one of the following predefined groups: patients examined by the gastroenterologist due to complaints with between 5 and 10 adenomas before the age of 50 years, 10-20 adenomas between the ages of 60 and 70 years or more than 20 adenomas over the age of 70 years, or patients with 10- 20 adenomas detected through population based screening aged between the ages of 55 and 75 years. Moreover, 7 patients without adenomas with multiple colorectal carcinomas were tested for *APC* mosaicism.

#### Rotterdam (EMC)

The Rotterdam cohort consists of patients who tested negative for pathogenic germline variants in adenomatous polyposis genes. Most patients tested for *APC* mosaicism (32/44) had ≥20 adenomas with or without colorectal cancer before the age of 70 years, 9/44 had ≥10 adenomas above the age of 60 years, one had >10 adenomas and >20 hyperplastic polyps at the age of 75 years and two had “multiple” adenomas of which one also had an adenoma of the papilla of Vater.

#### Groningen (UMCG)

The Groningen cohort also includes patients tested negative for pathogenic germline variants in adenomatous polyposis genes. *APC* mosaicism was tested whenever the detections of a mosaic variant would influence the surveillance guidelines for first-degree relatives. Therefore, this cohort includes patients with >20 adenomas and patients with 10-20 adenomas before the age of 55.

As previously categorized^5^, patients with a phenotype described as “FAP” (n=1) were considered to have >100 adenomas and “AFAP” (n=1) were considered to have 50-99 adenomas, and a description of “multiple adenomas” (n=4) was categorized as 30-49 adenomas and “some polyps” (n=1) as less than 10 adenomas. Gender information was not collected for the Rotterdam and Radboudumc cohort, resulting in 80 patients whose gender is “unknown”.

### Targeted Next-Generation Sequencing (NGS)

#### Leiden (LUMC)

DNA was extracted from Formalin-Fixed Paraffin-Embedded (FFPE) tissue blocks of preferably from 4 colorectal adenomas and/or carcinomas per patient and analyzed by targeted NGS as previously described.^16^ Briefly, a custom panel was used consisting of: *APC, MUTYH, POLE, POLD1, NTHL1, MLH1, MSH2, MSH6, PMS2, MSH3, BMPR1A, RNF43, PTEN, SMAD4, STK11, ENG, BRCA1, BRCA2, PALB2, TP53*. Ampliseq NGS libraries were prepared according to the manufacturer’s instructions. Sequencing was then performed on an Ion GeneStudio S5 Series sequencer (Thermofisher Scientific). Unaligned sequence reads were mapped against the human reference genome (hg19) using TMAP software and variants were called using Torrent Variant Caller.

Detected variants were classified by pathogenicity and compared between the lesions within each patient. Mosaicism was considered when all lesions shared an identical variant. Hybrid mosaicism was considered when an identical variant was detected in a subset of lesions. When mosaicism was detected in colorectal adenomas and/or carcinomas, leukocytes, buccal mucosa, and urine DNA was tested to determine the mosaic pattern throughout the body.

#### Rotterdam (EMC)

DNA from ≥2 FFPE tissues and *APC* was analyzed using a custom targeted NGS panel consisting of *APC* and *MUTYH*. Ampliseq NGS libraries were prepared according to the manufacturer’s instructions and sequenced on an Ion S5 XL system (Thermofisher Scientific). Sequencing data were analyzed using the Torrent variant caller (Thermofisher Scientific) and SeqPilot (JSI medical systems). Detected variants were classified according to pathogenicity and compared between the lesions within each patient.

#### Groningen (UMCG)

Samples from the UMCG were tested for *APC* at the Radboudumc using a custom NGS panel based on single-molecule molecular inversion probe (smMIP) enrichment. The probes covered all regions and intron-exon boundaries, as described previously.^17, 18^ Sequencing was performed on an Illumina NextSeq 500 or Novaseq 6000. Fastq files were analyzed using the SeqNext software package (JSI Medical Systems GmbH, Kippenheim, Germany). Consensus reads were generated, based on the single-molecule tag, and coding regions variants were called if present in ≥5% of all reads and ≥3 unique variant reads. Detected variants were classified by pathogenicity and compared between the lesions within each patient.

### Body Mass Index (BMI) and lifestyle data

For the LUMC cohort, BMI and lifestyle data were collected from patients’ medical records. In addition, a questionnaire about height, weight, smoking status, alcohol consumption, and medication use was mailed to 65 patients. Body Mass Index (BMI) was categorized as “underweight” with a BMI of ≤18.4, “healthy weight” with a BMI of 18.5-24.9, ‘overweight’ with a BMI 25.0-29.9 and “obese” with a BMI of ≥30.0. Smoking status and alcohol consumption were categorized as “never”, “former” and “current”, and pack-years (PY) of smoking were determined by the number of packs of cigarettes smoked per day multiplied by the number of years of smoking. In total, BMI was determined in 135 patients, smoking status in 229 patients, and alcohol consumption in 219 patients. Information on medication use was collected for approximately 60 patients.

### Statistical analysis

Statistical analyses were performed using IBM SPSS Statistics 25, and a p-value of <0.05 was considered significant. Independent t-tests, chi-square tests, and Fisher’s exact tests were used to assess phenotypic and lifestyle differences.

Multivariable logistic regression analysis was performed to determine associations between *APC* mosaicism and covariates of interest. Covariates included cumulative adenoma count (<10, 10-19, 20-49, 50-99, >100), age at diagnosis (<50, 50-59, 60-69, and >70), history of CRC (no, <50, 50-59 and >60), and the presence of extracolonic manifestations associated with FAP (yes, no). Results were reported as odds ratios, with 95% confidence intervals.

## Results

### *APC* mosaicism prevalence

This study included 541 patients who were tested for *APC* mosaicism at university medical centers in Leiden, Rotterdam, and Groningen using targeted NGS on DNA isolated from colorectal adenomas or carcinomas. A substantial proportion of patients (n=219) did not fall within the scope of the national colorectal cancer and polyposis testing guidelines; ≥10 adenomas before the age of 60 or with ≥20 adenomas before the age of 70.

A mosaic *APC* variant was detected in 9.4% (51/541) of patients, as summarized in Table 1. This detection rate was 14.3% (46/322) and 2.3% (5/219) in patients falling within and outside the scope of the national testing guidelines, respectively. Figure 1 shows a detection rate of 10% or more in patients with ≥20 adenomas before the age of 60 or ≥30 adenomas before the age of 70.

**Table 1.** Comparison of phenotypic characteristics between cases with and without *APC* mosaicism. Gender, adenoma count, age at first adenoma and CRC were significantly different between the two groups.

|  | **Total** | ***APC* mosaicism** | **No *APC* mosaicism** | ***p*-value** |
| --- | --- | --- | --- | --- |
| **Total** | 541 | 51 | 490 |  |
| **Gender – n (%)** |  |  |  |  |
| Male | 312 | 17 (33.3) | 295 (60.2) | 0.005* |
| Female | 149 | 20 (39.2) | 129 (26.3) |  |
| Unknown | 80 | 14 (27.5) | 66 (13.5) |  |
| **Adenoma count – n (%)** |  |  |  | <0.001* |
| 0 | 7<sup>#</sup> | 0 (0) | 7 (1.4) |  |
| 1-9 | 66 | 2 (3.9) | 64 (13.1) |  |
| 10-19 | 205 | 6 (11.8) | 199 (40.6) |  |
| 20-29 | 156 | 13 (25.5) | 143 (29.2) |  |
| 30-49 | 69 | 10 (19.6) | 59 (12.0) |  |
| 50-99 | 26 | 13 (25.5) | 13 (2.7) |  |
| >100 | 12 | 7 (13.7) | 5 (1.0) |  |
| **Age first adenoma – mean (min-max)** | 57.7 (17-84) | 44.1 (17-72) | 59.2 (22-84) | <0.001* |
| **CRC – n (%)** | 144 | 17 (33.3) | 127 (25.9) | 0.249 |
| **Age first CRC – mean (min-max)** | 57.0 (21-84) | 47.3 (25-69) | 58.4 (21-84) | <0.001* |
| **Extracolonic manifestations associated with FAP – n (%)** | 28 | 5 (9.8) | 23 (4.7) | 0.085 |

<sup>#</sup> patients without adenomas included because of multiple colorectal carcinomas

**Figure 1.**
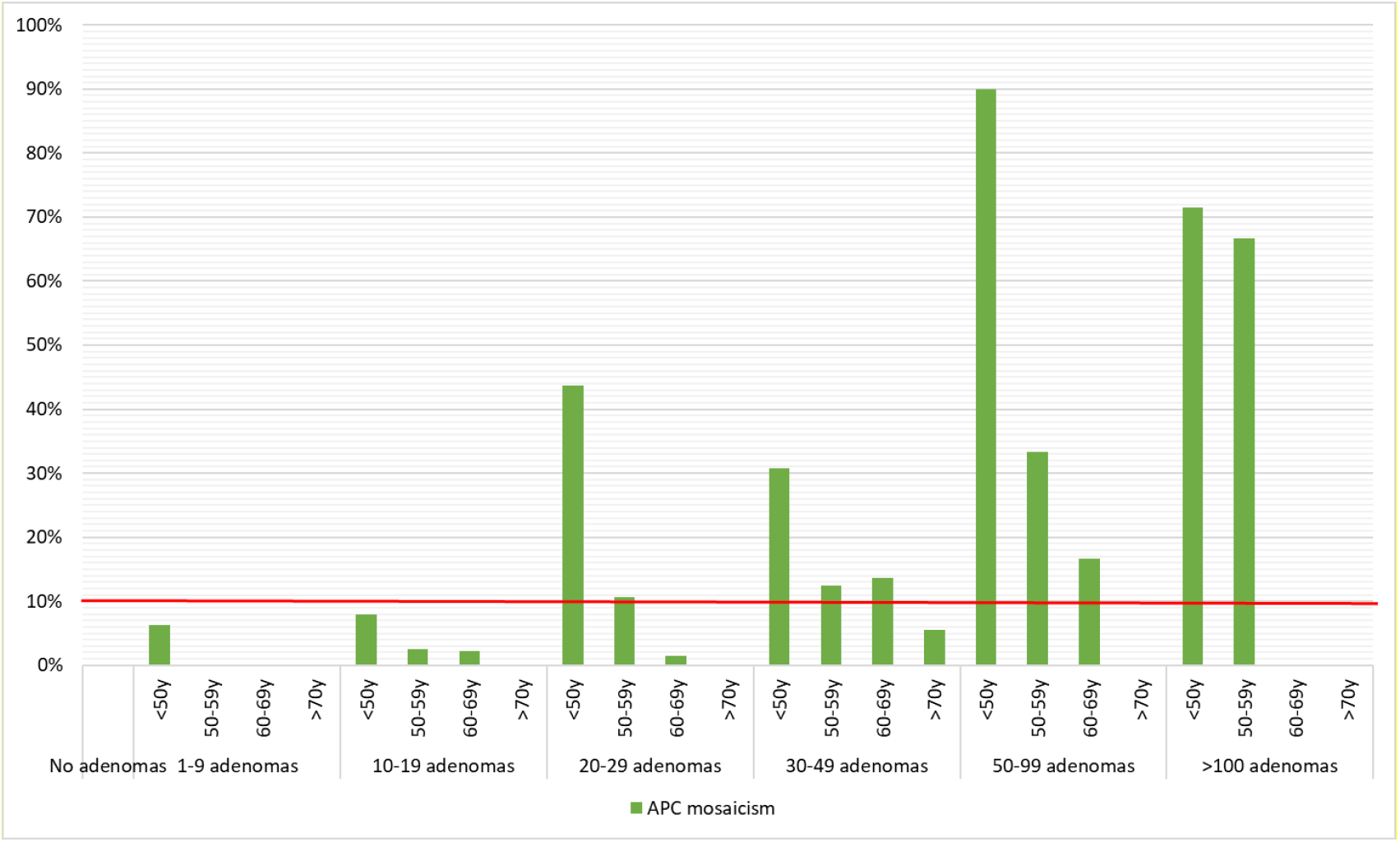
Detection rates of *APC* mosaicism subdivided in adenoma count groups and stratified for age at first adenoma.

In total, 103 patients with adenomas before the age of 50 years were analyzed with a detection rate of 28% (29/103). Clinical data of these patients are summarized in Supplemental Table 1. As also depicted in figure 1, the detection rate in patients with 1-10 adenomas before the age of 50 years is 5.7% (2/35). Moreover, no mosaicism was found in the seven patients without colorectal adenomas who were included because of multiple CRCs.

Mosaic patients have significantly more colorectal adenomas and develop the first adenoma at a younger age than non-mosaic patients. There was no significant difference in the prevalence of colorectal carcinomas (CRC), but when suffering from CRC, *APC* mosaic patients were diagnosed at a significantly younger age.

Multivariable logistic regression analysis in Table 2 supports this finding by showing significantly increased odds of finding a mosaic *APC* variant with higher adenoma counts 10-19: OR 5.0 [95% CI 0.5-46.3], 20-29: OR 20.9 [95% CI 2.4-179.4], 30-49: OR 23.9 [95% CI 2.6-221.0], 50-99 OR 126.9 [95% CI 13.5-1192.4], >100 OR 57.3 [95% CI 4.2-789.7] p-value <0.001) and at a younger age of adenoma diagnosis (<50: OR 35.5 [95% CI 4.1-304.5], 50-59: OR 5.6 [95% CI 0.6-50.0], 60-69 OR 2.5 [95% CI 0.3-21.3] p-value <0.001). A personal history of CRC or extracolonic manifestations associated with FAP did not affect the odds of detecting *APC* mosaicism.

**Table 2.** Multivariable logistic regression analysis. The odds of finding an *APC* mosaic case increase significantly upon adenoma count and a younger age of first adenoma

|  | n | OR (95% CI) | *p-value* |
| --- | --- | --- | --- |
| Adenoma count |  |  |  |
| 1-9 | 62 | Ref | <0.001* |
| 10-19 | 200 | 5.0 (0.5-46.3) |  |
| 20-29 | 144 | 20.9 (2.4-179.4) |  |
| 30-49 | 63 | 23.9 (2.6-221.0) |  |
| 50-99 | 22 | 126.9 (13.5-1192.4) |  |
| >100 | 9 | 57.3 (4.2-789.7) |  |
| Age first adenoma |  |  |  |
| >70 | 60 | Ref | <0.001* |
| 60-69 | 193 | 2.5 (0.3-21.3) |  |
| 50-59 | 155 | 5.6 (0.6-50.0) |  |
| <50 | 92 | 35.5 (4.1-304.5) |  |
| CRC (age first) |  |  |  |
| No | 374 | Ref | 0.881 |
| <50 | 26 | 0.9 (0.2-3.9) |  |
| 50-59 | 33 | 1.4 (0.3-7.1) |  |
| >60 | 67 | 1.7 (0.4-7.5) |  |
| Extracolonic FAP manifestations |  |  |  |
| No | 474 | Ref | 0.580 |
| Yes | 26 | 1.5 (0.3-7.0) |  |

### *APC* mosaicism characteristics

#### Normal tissues tested

As summarized in Supplemental Table 2, leukocyte, urine and/or buccal swab DNA was analyzed for the mosaic variant in 37 patients. Different patterns of *APC* mosaicism were detected, ranging from the mosaicism restricted to the colon (e.g., L ID 2) to extensive mosaicism throughout the entire body (e.g., L ID 13). A trend towards a higher number of adenomas and finding the mosaic variant in at least one other tissue tested was detected (mean of no other tissue: 35.5, mean at least one other tissue: 59.7). In addition, the mosaic *APC* variant was detected in normal colonic mucosa in 53% (10/19 tested) of patients.

#### Extracolonic phenotype

Thirty-four mosaic patients underwent esophagogastroduodenoscopy (EGD) and nine of them were diagnosed with gastric or duodenal polyps. Except for one patient with extensive mosaicism and osteoma and lipomas (L ID 13), no other patients presented with manifestations associated with FAP outside of the gastrointestinal tract.

#### Heritability

In none of the at least 24 children of 17 mosaic patients the mosaic variant was detected in leukocyte DNA. Of note, leukocyte, urine, and buccal swap DNA was analyzed in 12 of these 17 mosaic patients and showed restricted mosaicism to the colon in 10 patients. In the two mosaic patients with a more extensive mosaicism (L ID 486 and L ID 324), the variant allele frequency in leukocyte, urine and buccal swap was ≤1%.

In addition, semen was analyzed in two patients (L ID 117, 281) with a desire to have children. In one of these patients, the mosaic variant was detected with a variant allele frequency of 15% and 18% in duplicate testing.

### BMI and lifestyle (LUMC cohort only)

Table 3 shows that the non-mosaic group consists of significantly more males and smokers and is more frequently overweight or obese than the mosaic group. The distribution of never, former, and current alcohol consumers is comparable between the two groups, but the number of glasses per week is significantly higher in the non-mosaic group.

**Table 3.** Comparison of BMI and lifestyle data between patients with and without *APC* mosaicism.

|  | **Total** | **APC mosaicism** | **No APC mosaicism** | **p-value** |
| --- | --- | --- | --- | --- |
| **Gender – n (%)** | 462 | 38 | 424 |  |
| Male | 312 (67.5) | 17 (44.7) | 295 (69.6) | 0.005* |
| **BMI – n (%)** | 135 | 11 | 124 | 0.042* |
| Underweight | 2 | 0 (0.0) | 2 (1.6) |  |
| Healthy | 47 | 8 (72.7) | 39 (31.5) |  |
| Overweight | 61 | 3 (27.3) | 58 (46.8) |  |
| Obese | 25 | 0 (0.0) | 25 (20.2) |  |
| **BMI – mean (min-max)** | 26.8 (16.5-48.4) | 23.1 (19.2-26.9) | 27.2 (16.5-48.4) | 0.004* |
| **Smoking – n (%)** | 229 | 21 | 208 | 0.051 |
| Never | 68 | 11 (52.4) | 57 (27.4) |  |
| Former | 82 | 6 (28.6) | 76 (36.5) |  |
| Current | 79 | 4 (19.0) | 75 (36.1) |  |
| **Smoking PY – mean (min-max)** | 14.5 (0-73) | 4.2 (0-51) | 15.6 (0-73) | 0.016* |
| **Alcohol – n (%)** | 219 | 19 | 200 | 0.152 |
| Never | 38 | 6 (31.6) | 32 (16.0) |  |
| Former | 12 | 0 (0.0) | 12 (6.0) |  |
| Current | 169 | 13 (68.4) | 156 (78.0) |  |
| **Glasses/week – mean (min-max)** | 15.0 (0-168) | 4.2 (0-14) | 15.5 (0-90) | 0.001* |
| **Aspirin – n (%)** | 65 | 4 | 61 |  |
| Yes | 9 | 1 (25.0) | 8 (13.1) | 0.458 |
| **NSAIDs – n (%)** | 62 | 3 | 59 |  |
| Yes | 3 | 0 (0.0) | 3 (5.1) | 1.000 |
| **Antihypertensive – n (%)** | 64 | 3 | 61 |  |
| Yes | 28 | 3 (100.0) | 25 (41.0) | 0.079 |

### Hybrid mosaicism prevalence and phenotype (LUMC cohort only)

Ninety-seven patients (21%) were identified with so-called hybrid mosaicism, multiple but not all lesions sharing the same *APC* variant. The results are summarized in Supplemental Table 3. Notably, 37 of the 97 hybrid mosaic patients have a recurrent *APC* variant that fits the colibactin-associated mutational signature as described previously.^16^

Table 4 shows that hybrid mosaic cases were significantly different from the *APC* mosaic cases in terms of gender, adenoma count, and age at first adenoma. However, hybrid mosaics were phenotypically comparable to the non-mosaic group. Therefore, all descriptions and analyses included hybrid mosaic cases as non-mosaic cases.

**Table 4.**
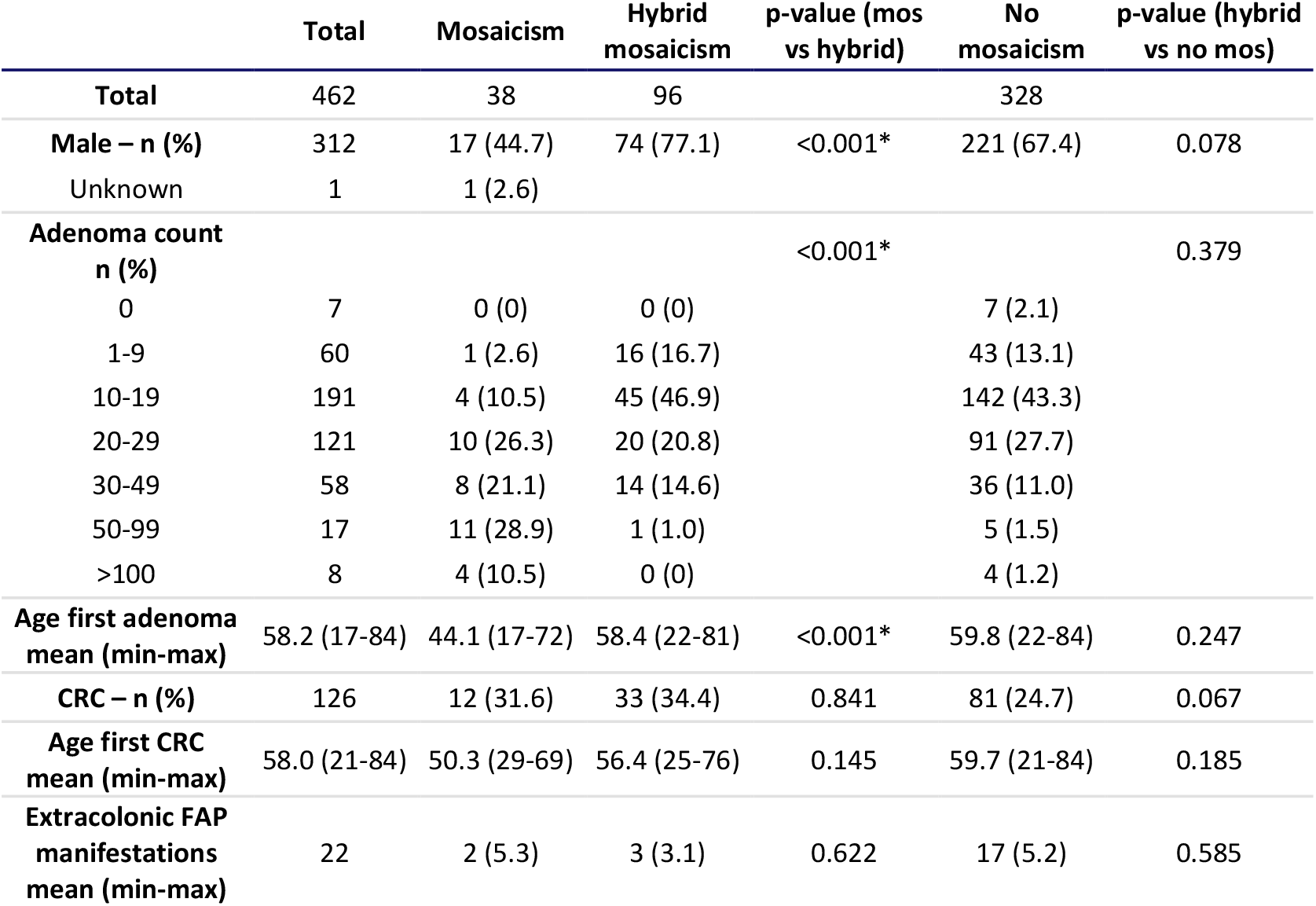
Phenotypic comparison of hybrid cases with mosaic cases and no mosaic cases. Hybrid mosaic cases are more comparable with cases without *APC* mosaicism.

|  | Total | Mosaicism | Hybrid mosaicism | p-value (mos vs hybrid) | No mosaicism | p-value (hybrid vs no mos) |
| --- | --- | --- | --- | --- | --- | --- |
| **Total** | 462 | 38 | 96 |  | 328 |  |
| **Male – n (%)** | 312 | 17 (44.7) | 74 (77.1) | <0.001* | 221 (67.4) | 0.078 |
| Unknown | 1 | 1 (2.6) |  |  |  |  |
| **Adenoma count n (%)** |  |  |  | <0.001* |  | 0.379 |
| 0 | 7 | 0 (0) | 0 (0) |  | 7 (2.1) |  |
| 1-9 | 60 | 1 (2.6) | 16 (16.7) |  | 43 (13.1) |  |
| 10-19 | 191 | 4 (10.5) | 45 (46.9) |  | 142 (43.3) |  |
| 20-29 | 121 | 10 (26.3) | 20 (20.8) |  | 91 (27.7) |  |
| 30-49 | 58 | 8 (21.1) | 14 (14.6) |  | 36 (11.0) |  |
| 50-99 | 17 | 11 (28.9) | 1 (1.0) |  | 5 (1.5) |  |
| >100 | 8 | 4 (10.5) | 0 (0) |  | 4 (1.2) |  |
| **Age first adenoma mean (min-max)** | 58.2 (17-84) | 44.1 (17-72) | 58.4 (22-81) | <0.001* | 59.8 (22-84) | 0.247 |
| **CRC – n (%)** | 126 | 12 (31.6) | 33 (34.4) | 0.841 | 81 (24.7) | 0.067 |
| **Age first CRC mean (min-max)** | 58.0 (21-84) | 50.3 (29-69) | 56.4 (25-76) | 0.145 | 59.7 (21-84) | 0.185 |
| **Extracolonic FAP manifestations mean (min-max)** | 22 | 2 (5.3) | 3 (3.1) | 0.622 | 17 (5.2) | 0.585 |

## Discussion

This study reported on 541 patients, with multiple colorectal adenomas or carcinomas who were tested for *APC* mosaicism in Leiden, Rotterdam, and Groningen. With 51 mosaic patients, a detection rate of 9.4% was found among this selected group. This rate was 14.3% (46/322) for patients falling within the scope of the national Polyposis and Colorectal Cancer Guidelines for testing for germline variants; ≥10 adenomas before the age of 60 years and ≥20 adenomas before the age of 70 years^15^ and 2.3% (5/219) in patients outside the scope. These rates were significantly lower than those in the study by Jansen et al.^19^, which found *APC* mosaicism in patients with >20 adenomas (9/18; 50%), although this study was small. However, the detection rate in the current cohort was 19% (42/219) in patients with >20 adenomas before the age of 70, which is comparable to another previously performed study.^12^

As expected, the prevalence of *APC* mosaicism increased with the number of adenomas and with younger age at adenoma diagnosis. Other factors, such as CRC diagnosis or FAP-associated extracolonic tumors, did not influence the odds of finding *APC* mosaicism.

Most mosaic patients presented with ≥10 adenomas before the age of 60 or ≥20 adenomas before the age of 70 and, therefore, fall within the national Hereditary Polyposis Guidelines, which are comparable to the National Comprehensive Cancer Network guideline for genetic/familial colorectal cancer.^14, 15^ More specifically, *APC* mosaicism rates were ≥10% in patients with ≥20 adenomas before the age of 60, or ≥30 adenomas before the age of 70. Based on these data, we would recommend *APC* mosaicism testing at least in all patients tested negative for pathogenic germline variants with (1) ≥20 adenomas before the age of 60 years or (2) ≥30 adenomas before the age of 70 years.

This cohort showed different patterns of *APC* mosaicism, ranging from extensive mosaicism throughout the entire body to mosaicism restricted to the colon. A spectrum of colorectal phenotypes was also observed; mosaic patients with e.g., ten adenomas at 59 years of age as well as extensive adenomatous polyposis. This broad spectrum complicates universal guidelines for endoscopic surveillance. We suggest that the colonoscopy frequency for *APC* mosaic patients should in principle be comparable to the (A)FAP guidelines^21^; every one or two years. In case a patient has a mild phenotype with effective polypectomy, the frequency of colonoscopy could be re-evaluated to, for example, once every three years.

In addition, duodenal or gastric adenomas were detected in 9 out of 34 mosaic patients who underwent an EGD. Therefore, based on our data, we would suggest an EGD at least once at the age of 35, with the frequency of follow-up determined by the findings of the first EGD.

Besides the consequences for the index patient, the identification of *APC* mosaicism is also relevant for family members. According to the national surveillance guidelines, parents, siblings, and children of patients with unexplained polyposis are offered regular colonoscopies.^15^ Since mosaicism occurs during embryogenesis and is, therefore, an isolated case in a family, asymptomatic parents and siblings can be reassured, and no surveillance colonoscopies are required.

Germline testing of at least 24 children of 17 mosaic patients showed that the variant was not transmitted to their offspring. This may suggest, as previously described^19^, that the likelihood of inheritance is low. Notably, DNA from leukocytes, urine, and buccal swabs was analyzed in 12 of these 17 patients. In ten of them, the analysis showed that the mosaicism was confined to the colon. In the two mosaic patients with a more extensive mosaicism (L ID 486 and L ID 324), the variant allele frequency in leukocyte, urine and buccal swap was ≤1%. Although more research is needed, this low variant allele frequency may also indicate a low likelihood of inheritance. In addition, a mosaic variant was detected in the semen DNA of another patient who wanted to have children. Therefore, transmission cannot be excluded, and we still recommend germline testing in children aged above 12 years.

Six mosaic patients would have been missed using our proposed testing guidelines (L ID 10, 19, 31 and 361, R ID 43, G ID 35). One of these patients (L ID 361) has segmental polyposis, with most adenomas located on the left side of the colon. In this case, the mosaicism is probably restricted to the left side of the colon, which explains the milder phenotype. Another patient (R ID 43) was diagnosed with both colorectal and duodenal adenomas.

Two other patients (L ID 10 and 19) had mosaic variants that fit the mutational signature associated with colibactin.^16, 22, 23^ The development of adenomas is likely due to the presence of colibactin and may not be due to mosaicism. Therefore, these patients are unlikely to develop as many colorectal adenomas as ‘true’ mosaic patients. Further research on the association between colibactin, adenoma development, and possible prevention or eradication is needed to gain more insight and provide appropriate screening guidelines.

Although most hybrid mosaic cases had a recurrent *APC* variant that matched the colibactin mutational signature, other explanations include clonal relationship and contamination, such as combining two adenomas being grouped as one during colonoscopy or analysis. Since there is no universal explanation that fits all hybrid cases, a case-by-case evaluation is required. However, phenotypically hybrid cases are comparable to non-mosaic cases and should generally be treated as such.

The non-mosaic group had a significantly higher BMI, were more likely to be former or current smokers, and drank more glasses of alcohol per week than the mosaic patients. These findings suggest that these modifiable risk factors may have contributed to the development of colorectal adenomas in the non-mosaic group, as previously proposed for colorectal cancer.^24, 25^

In conclusion, we recommend testing for *APC* mosaicism in all patients with (1) ≥20 adenomas before the age of 60 years, or (2) ≥30 adenomas before the age of 70 years. Like FAP patients, we suggest that patients with *APC* mosaicism should be offered regular colonoscopy screening and esophagogastroduodenoscopy from the age of 35. The frequency of additional endoscopies should depend on the findings. Furthermore, despite the low chance of transmission, we recommend considering germline testing in children of mosaic cases.

## Data Availability

All data produced in the present study are available upon reasonable request to the authors

## Acknowledgements

The authors acknowledge the technicians of the Molecular Diagnostics unit of the department of Pathology Leiden University Medical Center for their help with the analyses.

